# National Strategy to Reduce Cervical Cancer Incidence and Mortality: Are Health Facilities Complying? Case of Kilimanjaro Region

**DOI:** 10.1101/2025.11.26.25341113

**Authors:** Paschal Yuda, Anthony Kavindi, Gasper Mpehongwa, Declare L. Mushi, Joackim Kessy

**Affiliations:** School of Public Health, KCMC University, P.O. Box 2240, Kilimanjaro Tanzania

**Keywords:** Cervical cancer screening, Secondary prevention Gaps, Health facility Readiness and compliance in Tanzania

## Abstract

**Background:** Cervical cancer remains the leading cause of cancer-related deaths among women in Tanzania despite being largely preventable. In response, the Ministry of Health developed the Tanzania Cervical Cancer Prevention and Control Strategic Plan (2020–2024), emphasizing secondary prevention strategies such as screening and pre-cancer treatment by health facilities across the country. However, by the year 2020, only 9% out of 6,447 health facilities were providing screening and treatment services in Tanzania.

**Methods:** A cross-sectional quantitative study was conducted in the Kilimanjaro Region. Ten (10) health facilities offering cervical cancer screening services. A stratified sampling method was employed. Data was collected using the CECAP scorecard checklist. Data was analyzed using STATA 18, and facility performance was assessed based on readiness and guideline compliance thresholds.

**Results:** Among 10 facilities that were assessed, only 60% were structurally ready to deliver quality cervical cancer screening services, and just 30% complied with national screening guidelines. Gaps were observed in the availability of treatment equipment, trained healthcare providers, follow-up mechanisms, and inadequate community outreach services. Out of 462 health facilities in the region, only 41(8.9%) facilities offer cervical cancer screening services, indicating major gaps in service availability and coverage limitations.

**Conclusion:** The study reveals gaps in cervical cancer facility readiness and compliance with national guidelines in the Kilimanjaro region. Despite cervical cancer services’ availability, service integration, systemic, and operational barriers persist. Targeted investments in infrastructure, training, monitoring, and community engagement are essential to accelerate progress towards the national cervical cancer prevention goal.

## Introduction

Cervical cancer remains a significant global public health challenge, with an estimated 660,000 new cases and 350,000 deaths in 2022 (1). In Europe, cervical cancer incidence is lower compared to Sab-Saharan Africa (SSA), whereas SSA faces concerning rates, with an incidence of 29.79 per 100,000 people (2). In Tanzania, a study reported a cervical cancer rate of 4% among women attending a gynecological clinic, with just 6.5% having received screening (3).

Cervical cancer is the fourth most frequent malignancy in women worldwide, and 85% of deaths occur in low- and middle-income nations (4). In Africa, cervical cancer is the primary cause of cancer-related mortality in women, with incident rates seven to ten times greater than High Middle-Income Countries (HMIC), SSA has the world’s highest incidence rates (5). Annually, 90,000 new cases of cervical cancer are diagnosed, with 57,000 deaths, and the disease burden has increased in several countries over the last few decades (6). The highest cervical Human Papillomavirus (HPV) incidence rates in SSA contribute to this status. The frequency is above the global average of 11%. East Africa has a 33% HPV prevalence, followed by West Africa (29%) and Southern Africa (17%) (6). The difference in prevalence is reported as early sexual debut, especially in East Africa (7), also low condom use in East and West Africa due to restricted access or gender norms worsens transmission. Examples in Southern Africa have a greater condom use rate of 80% which lowers HPV transmission and a well-structured health system compared to East and West Africa (8).

In response to these alarming numbers of cervical cancer, WHO established prevention strategies which are primary (HPV vaccination), secondary (screening and treatment of precancerous lesions), and tertiary (treatment of invasive cancer), and adopted by the Tanzanian Ministry of health (9,10). Over the recent decade, there has been a substantial attempt to develop national immunization programs, two countries stand out, Burundi and Rwanda, where adolescent girls receive vaccinations exceeding 89% coverage and have previously shown a decrease in HPV prevalence after implementation (6). In SSA, the availability and utilization of cervical cancer screening services vary widely across countries. According to a study by Jedy-Agba *et al*, conducted in Sab-Saharan countries only 11 out of 47 countries in the region have established national cervical cancer screening programs (11). In many countries, opportunistic screening approaches are more common, leading to uneven coverage and disparities in access to screening services (11).

In Tanzania, 18.8 million women aged 15 years and older may have cervical cancer. A total of 10,241 women are diagnosed with cervical cancer annually in the country, out of which 63.7% die from the disease (12). The disease is the primary cause of cancer-related death and imposes a significant cost on families and the healthcare system. Structural constraints, including shortages of skilled providers, insufficient screening rooms and equipment, restricted funding, and sociocultural obstacles, contribute to chronically poor screening coverage (13,14). A recent study further emphasized the influence of cultural views and insufficient health literacy on women’s health-seeking habits, further reducing uptake (13).

In response to this persistent burden, Tanzania’s Ministry of Health, Community Development, Gender, Elderly, and Children (MoHCDGEC) formulated the Tanzania Cervical Cancer Prevention and Control Strategic Plan (2020–2024). This national strategy intends to reduce cervical cancer incidence and mortality by expanding access to high-quality screening and treatment services, with a stated goal of increasing the proportion of health facilities providing these services from 9% to 50% by 2024 (10). The approach emphasizes the adoption of cost-effective procedures like Visual Inspection with Acetic Acid (VIA), along with prompt treatment using cryotherapy or thermoablation, in accordance with the World Health Organization’s (WHO) global appeal to eliminate cervical cancer (9). Despite these efforts, inequalities in service availability and provider adherence to national guidelines remain, and monitoring of facility-level implementation has been minimal (15–18).

Despite the Kilimanjaro region’s established healthcare infrastructure and strong participation in cervical cancer prevention activities, uptake remains inadequate. A recent study in Moshi Municipality found that poor participation rates were due to inadequate structural preparation, a lack of qualified staff, and uneven adherence to screening recommendations (19). These challenges highlight the importance of assessing how well regional facilities are equipped and whether national and WHO guidelines are being implemented as intended.

Therefore, this study intends to evaluate the structural readiness of health facilities and their compliance with national guidelines for cervical cancer screening in the Kilimanjaro region under the 2020-2024 national strategic plan. The assessment’s findings, which identify specific gaps and strengths in the current service delivery system, will provide policymakers and health managers with actionable evidence to strengthen the cervical cancer prevention program and accelerate progress toward national and global elimination goals.

## Methods

### Study Design and Setting

This study used a cross-sectional study design and utilized a quantitative approach to provide a comprehensive understanding of the facility readiness and compliance to national guidelines for cervical cancer in the Kilimanjaro region from 01/07/2025 to 31/07/2025. Kilimanjaro region, located in the northeastern part of Tanzania. The Region is purposefully chosen for this study due to its significant population size, established healthcare infrastructure, including the zonal referral hospital of KCMC, and ongoing efforts including provision of cervical cancer screening, integration of screening services to other with other related health services and capacity building to healthcare providers in cervical cancer prevention. In the Kilimanjaro region, women account for around 51.3% of the overall population, with those aged 30 to 49 accounting for approximately 41.0% of all women (20). The Kilimanjaro region is divided into seven administrative district councils: Moshi Municipal, Moshi District, Hai, Rombo, Mwanga, Same, and Siha. Each district varies in terms of population density, healthcare facilities, and access to health services. Kilimanjaro Region is actively engaged in various cervical cancer prevention and control initiatives, aligned with Tanzania’s national strategy to combat the disease. The region offers cervical cancer screening, such as Visual Inspection with Acetic Acid (VIA), Pap smear tests, and Human Papillomavirus (HPV) DNA testing. Screening services are integrated into routine reproductive health services, and community outreach programs are conducted to raise awareness and encourage women to participate in regular screening. The Kilimanjaro region has a total of 462 health facilities both private and public of all levels, among them only 41 (8.9%) health facilities offer cervical cancer screening cervices.

### Study Population and Sample Size

The study population consisted of dispensary, health centers, district-level and referral hospitals health facilities that provided screening services in the region.

### Sampling Technique

Stratified sampling was applied to select health facilities in the study. Kilimanjaro has 7 district councils (Moshi MC, Moshi DC, Hai, Rombo, Siha, Same, and Mwanga) with 462 health facilities and only 41 facilities offering cervical cancer screening which are Moshi MC 6, Moshi DC 11, Hai 8, Rombo 6, Siha 3, Same 4, and Mwanga 3 facilities of different levels.

Among 41 facilities, referral or specialized hospitals are 3, district-level hospitals (13), health centers (24) and only 1 dispensary. To ensure coverage across all levels, the sample contained 10 facilities chosen from each stratum, guided by the proportion of facilities in each category. The study covered 2 referral hospitals which were purposively selected. Mawenzi regional referral hospital as it serves average number of about 300 out-patients daily and it implement cervical cancer screening. The second one is KCMC zonal referral with well-established clinics that provide screening, diagnosis, treatment, and follow-up services, also serves as a training and research facility, making it excellent for comprehensive examination. We excluded Kibong’oto specialized hospital as it deals more with infectious diseases.

Three district-level hospitals, namely Huruma Hospital, Machame Lutheran Hospital and Same District Hospital were purposively selected, two private and one public. Huruma Hospital in Rombo DC (Private) is the key provider of cervical cancer services in the eastern part of Kilimanjaro, has participated in reproductive health outreach and screening activities. Machame Lutheran Hospital, located in Hai DC (private), is a well-established hospital with good infrastructure, serving a large population of approximately 150,000 people in the surrounding area, and is actively engaged in women’s health services. Same District Hospital from Same DC is a government district hospital that provides both screening and treatment, and serves the eastern part of the region’s population.

Four health centers were randomly selected from areas where referral and district hospitals were selected: Mwanga health center from Mwanga DC, Siha health center from Siha DC, Himo and Kahe health centers from Moshi DC. Chama cha Uzazi na Malezi Bora dispensary, as the only dispensary offering cervical cancer screening in Kilimanjaro was also purposively selected.

### Study Variables

#### Dependent Variables

**S**tructural readiness was measured using a CECAP scorecard tool from MoH and it was further categorized <70 as low quality and ≥70 as good quality and health facilities’ compliance with national cervical cancer screening guidelines was also measured using a CECAP scorecard tool from MoH and it was further categorized <70 as low compliance and ≥70 as good compliance.

#### Independent Variables

Healthcare facility type, availability of VIA, cryotherapy, thermoablation, private area for ccs and cryotherapy, private area for ccs and thermoablation, examination table, light source, specula, acetic acid, jik, methods for specula sterilization, carbon-dioxide gas, trained providers for VIA, cryotherapy and thermoablation, PITC/ CTC integration, FP services integration, maternal services integration, SRH services integration, STI services integration, Inpatient services integration, integration guidelines, registers/ logbooks, data discrepancies, cryotherapy follow-up, thermoablation follow-up, referral follow-up, client card, IEC materials, community education and facility-based education.

### Data Collection Tools and Methods

Data collection tool used was a CECAP score card checklist. The CECAP score card is a checklist structured for facility-based assessment designed for monitoring and evaluation of the implementation of cervical cancer services within healthcare facilities. Its primary focus is on screening, integrating screening with other health services, follow-up and health promotion activities. The tool allows program implementers, facility administrators, and health system evaluators to systematically evaluate the availability, accessibility, and quality of these key services. Interviews with a health worker using CECAP scorecard on a facility visit for facility assessment was also conducted.

### Statistical Analysis

The statistical analysis was conducted using STATA version 18.0 (STATA Corp, College Station, TX). Data from health facilities were analyzed using the CECAP scorecard. A composite score ranging from 0 to 16, which was converted into a percentage, was assigned based on the availability of essential infrastructure, qualified personnel, equipment, and established procedures. Facilities with a score of ≥70% on the CECAP score card were categorized as offering high-quality cervical cancer services, and those with scores below 70% were categorized as having low-quality service for cervical cancer secondary prevention, while facilities scoring ≥70% were categorized as having high-quality of service cervical cancer secondary prevention. Descriptive statistics was used to summarize facility-level indicators and proportion of facilities categorized as high or low quality. Frequency and percentages were reported.

On health facilities’ compliance with national cervical cancer screening guidelines, A CECAP core card checklist was used to collect data on proper documentation, follow-up, counseling, service frequency, VIA use, and IEC materials were the key indicators evaluated and use a composite score ranging from 0 to 16 which was converted into percentage to assess compliance level. Facilities scoring ≥70% were considered as having high compliance level, while those scoring below 70% were categorized as having low compliance level. Results were summarized using frequencies and percentages to show distribution of compliance scores and key service components across assessed facilities.

## Results

### Distribution of Health Facilities

The assessment included 10 facilities across facility levels in the Kilimanjaro region. At dispensary level we had 1 facility, 4 health centers, 3 district-level hospitals and 2 referral hospitals. Geographically the facilities were distributed across all 7 districts of Kilimanjaro region, Moshi MC with highest presentation of 3 facilities, Moshi DC with 2 facilities while the rest of districts have only 1 facility each.

**Table 1:** Health facility distribution.

|  | n=10 | Percent |
| --- | --- | --- |
| **Facility level** |  |  |
| Dispensary | 1 | 10.0 |
| District level | 3 | 30.0 |
| Health center | 4 | 40.0 |
| Referral hospital | 2 | 20.0 |
| **District councils** |  |  |
| Hai DC | 1 | 10.0 |
| Moshi DC | 2 | 20.0 |
| Moshi MC | 3 | 30.0 |
| Mwanga DC | 1 | 10.0 |
| Rombo DC | 1 | 10.0 |
| Same DC | 1 | 10.0 |
| Siha DC | 1 | 10.0 |

### Health Facility Structural Readiness

Assessment of structural readiness of selected facilities offering cervical cancer screening services in Kilimanjaro Regions revealed that 60% (6 out of 10) met the minimum readiness threshold, while 40% (4 out of 10) were low readiness according to readiness index cutoff of 70% for facility to manage cervical cancer (Fig 1).

**Fig 1:**
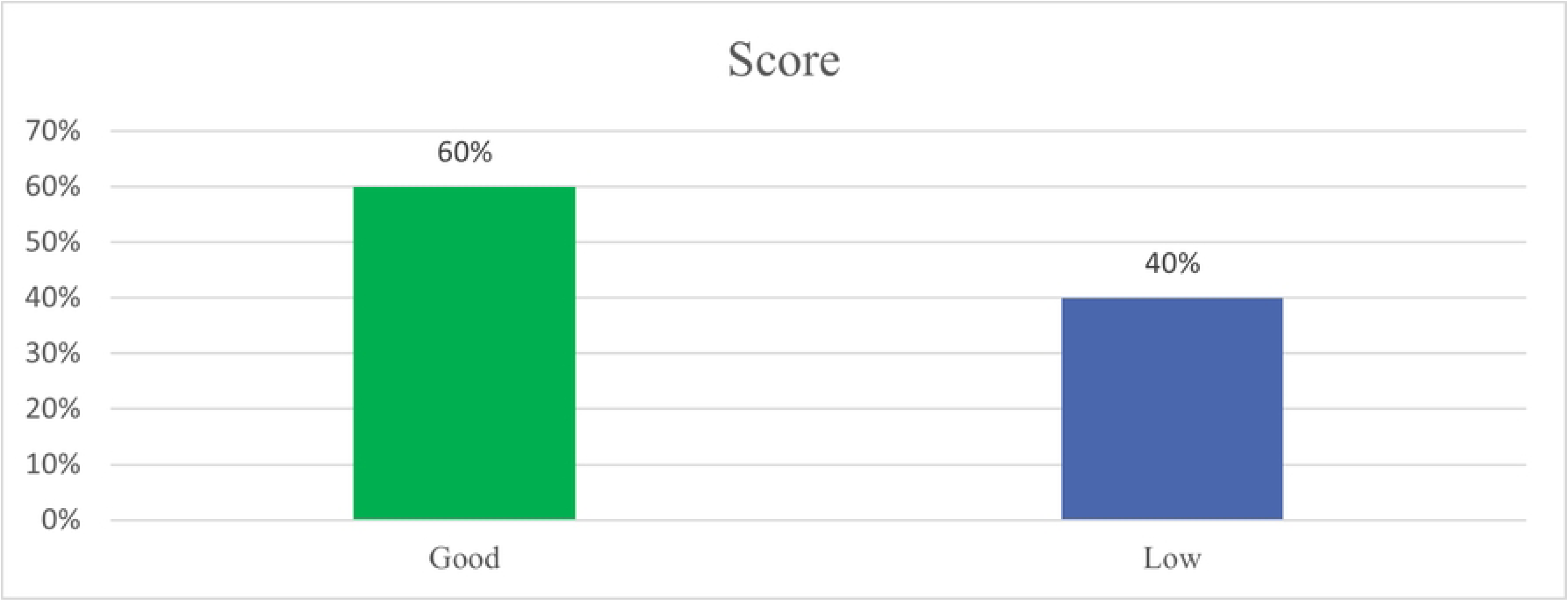
Structural readiness.

### CECAP Score Distribution

CECAP score distribution among assessed health facilities revealed significant variability in structural readiness. The score ranged from 3 to 16 with higher scores indicating better readiness. One facility scored low as 3 (19%) indicating poor in readiness. Additional 2 facilities scored 6 (38%) and 7 (44%) which is below readiness level. One facility scored 11 (69%) indicating moderate in readiness. Two facilities scored 12 (75%) while 3 facilities scored 13 (81%) suggesting good structural readiness and only one facility scored highest possible score of 16 (100%) indicting full structural readiness to offer high quality secondary prevention services for cervical cancer (Fig 2).

**Fig 2:**
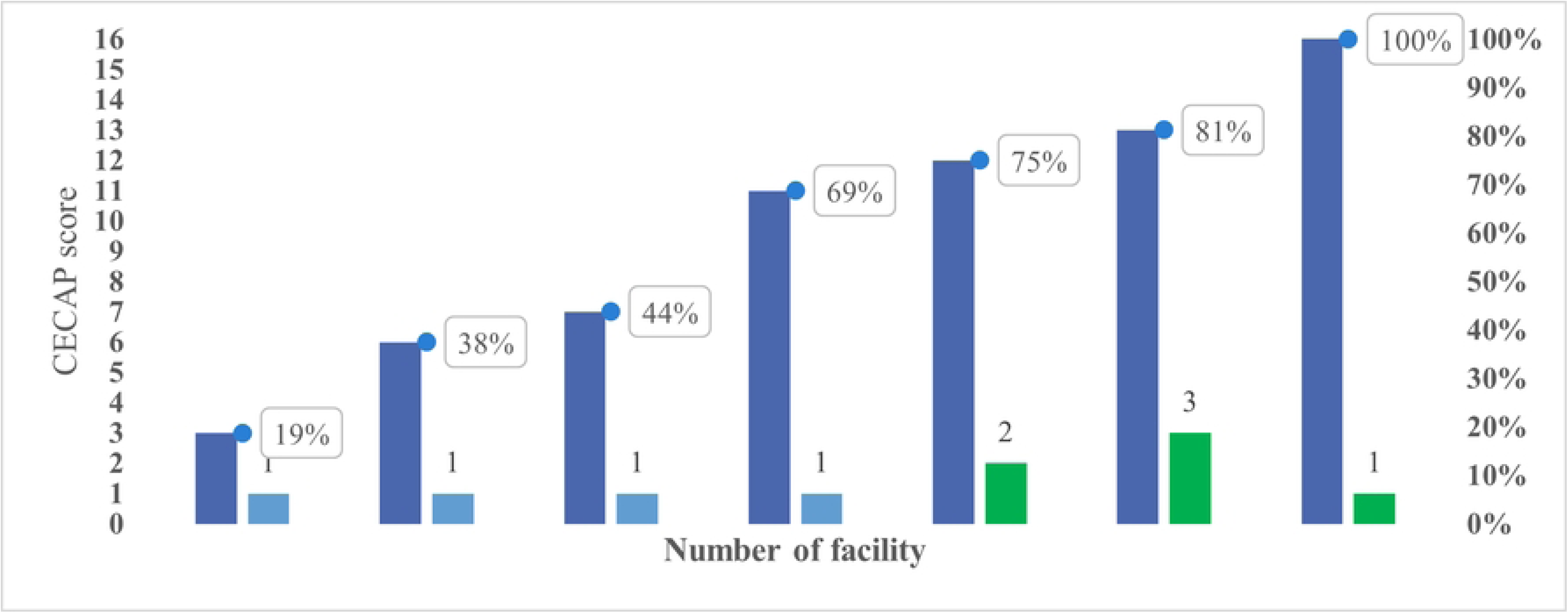
CECAP score distribution.

### Structural Readiness Scores

The findings from the assessment revealed that 90% (9 out of 10) facilities offer VIA, and 90% (9) facilities also have acetic acid available. However, only 50% (5) of facilities reported having cryotherapy equipment, and just 30% (3) of facilities offer thermoablation services. Private area for VIA and cryotherapy is available in 80% (8) facilities, but only 30% (3) have a private area for VIA and thermoablation.

Only 40% (4 out of 10) facilities have examination tables, and 90% (9) facilities have enough specula. Light sources are available and functional in 70% (7) of facilities. For infection prevention, 80% (8) of facilities had decontamination supplies, and all 100% (10) facilities reported having methods for specula sterilization. Adequate carbon dioxide (CO₂) for cryotherapy is available in only 50% (5) of facilities.

In terms of trained service providers, 80% (8) of facilities have VIA-trained providers, 40% (4) have providers trained for both VIA and cryotherapy, and only 30% (3) of facilities have providers trained for both VIA and thermoablation (Fig 3).

**Fig 3:**
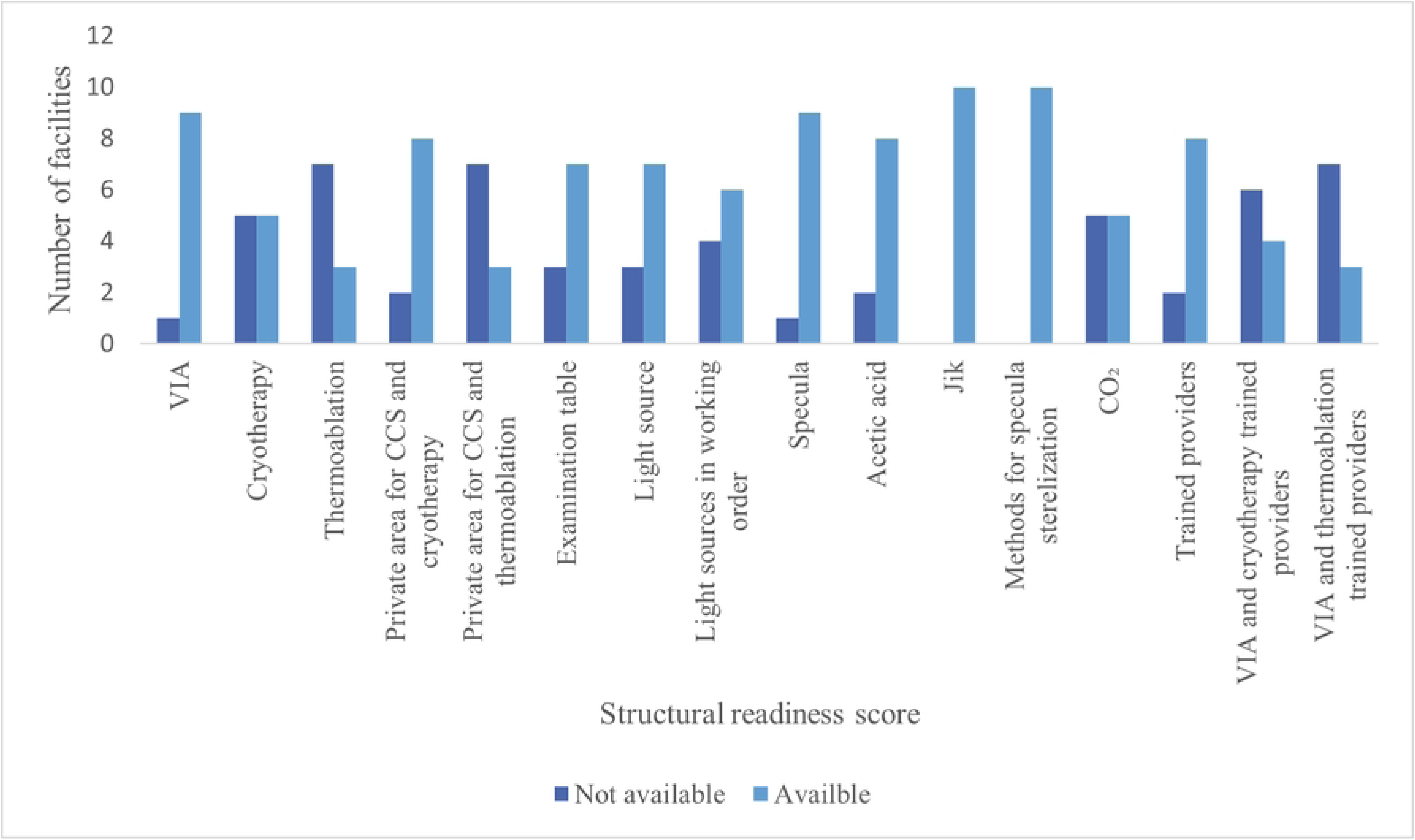
Structural readiness scores.

**Fig 4:**
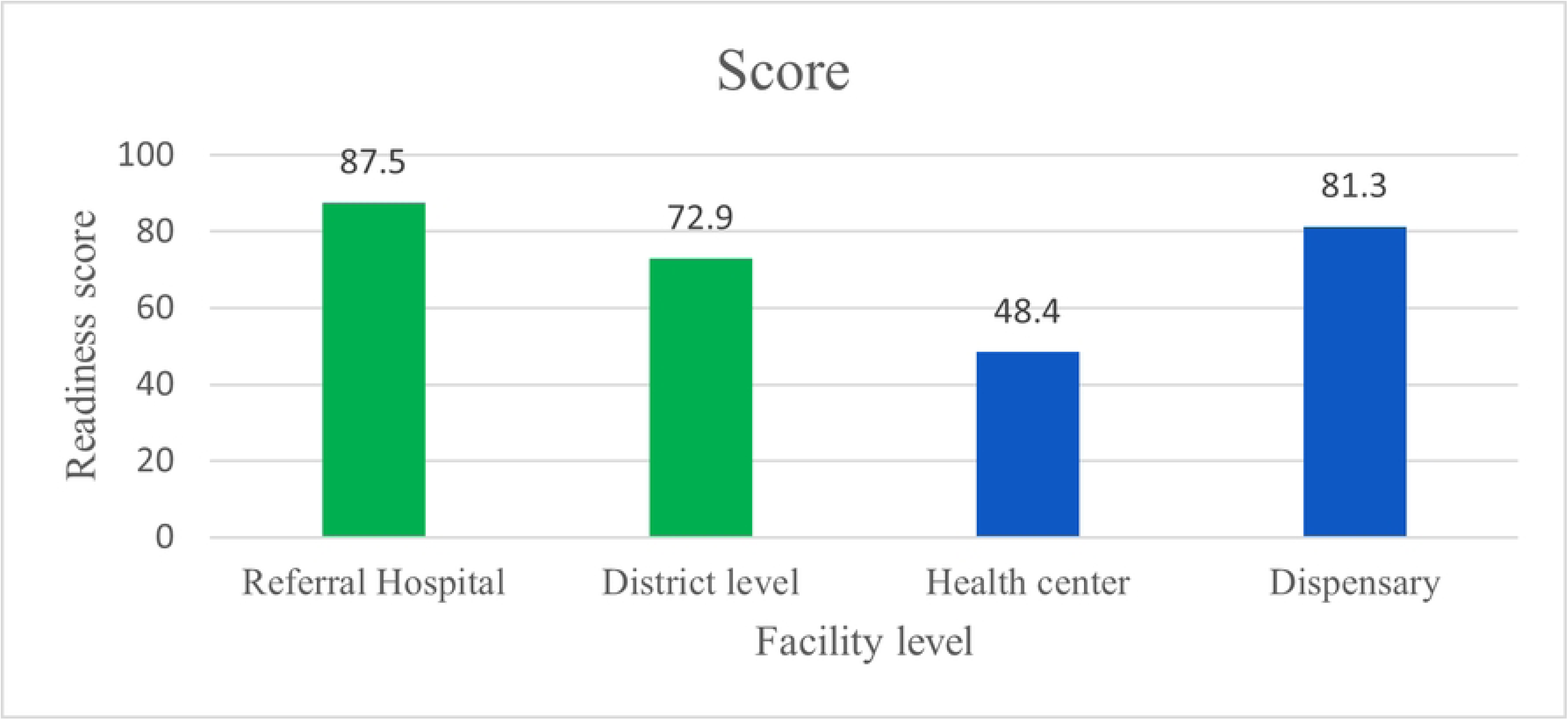
Facility-level structural readiness.

### Facility-Level Structural Readiness

Cervical cancer secondary prevention readiness varied notably across facility levels. Referral hospitals had the highest readiness score at an average of 87.5%, followed by the dispensary at 81.3%, and district level hospitals at 72.9%, all which met the readiness cutoff of ≥70%. In contrast, health centers had lowest score at 48.4%, indicating inadequate preparedness to deliver high-quality secondary prevention for cervical cancer services.

### Health Facilities’ Compliance with National Cervical Cancer Screening Guidelines

The assessment of health facilities’ compliance with national guidelines for cervical cancer screening revealed that the majority of health facilities demonstrate low compliance. 70% (7 out of 10) facilities had low compliance, while 30% (3 out 10) met the criteria for good compliance with the national guidelines by scoring above 70% on the CECAP score card (Fig 5).

**Fig 5:**
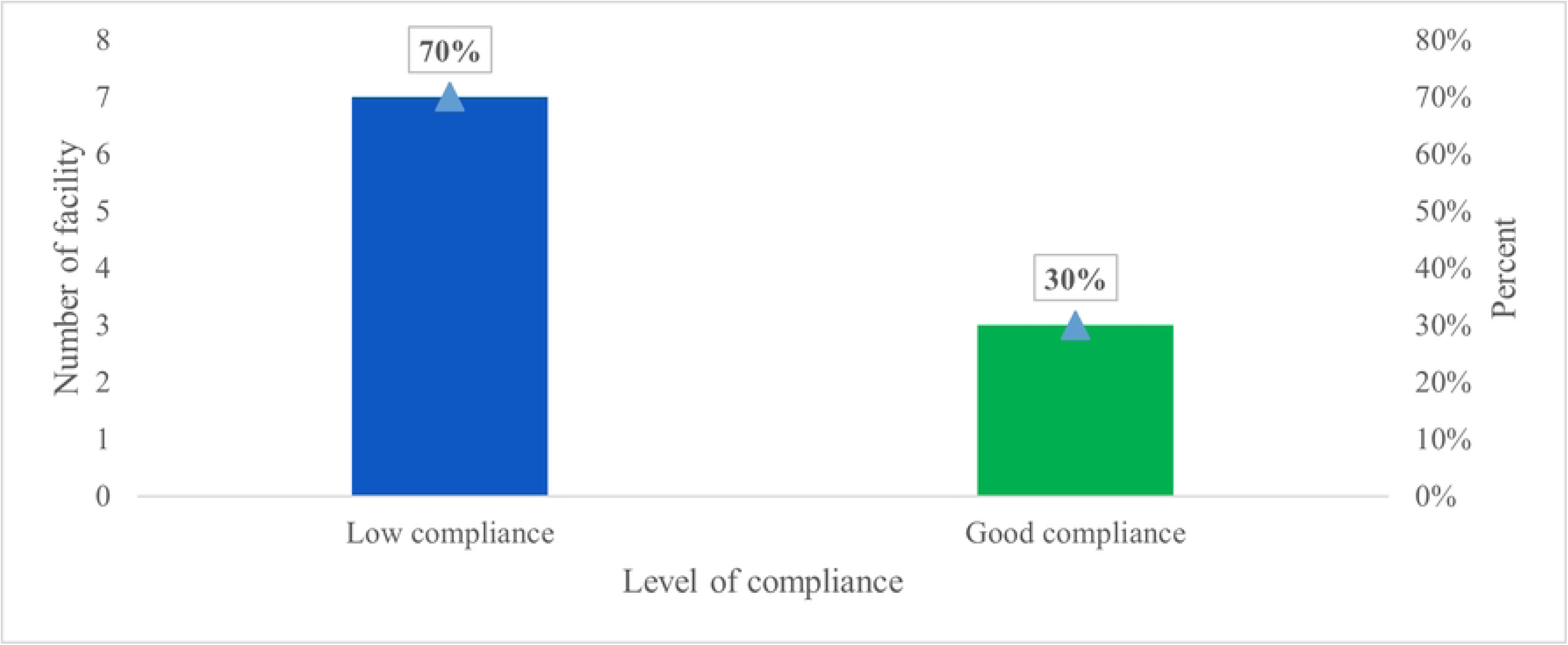
Compliance level.

### Compliance Score

The compliance scores of health facilities with the national cervical cancer screening guidelines varied significantly, ranging from a score of 0 to 13 as one facility score 0 (0%) indicating no compliance at all. The scores of 8,9 and 10 were recorded in 3 different facilities with corresponding compliance percentages of 50%, 56.3% and 62.5% respectively. 2 facilities scored 11 (68.8%) and 12 (75%), while the remaining 2 facilities achieved the highest score of 13 (81.3%). Although few facilities demonstrated relatively high compliance, the overall distribution shows that most facilities fall below the optimal compliance threshold (Fig 6).

**Fig 6:**
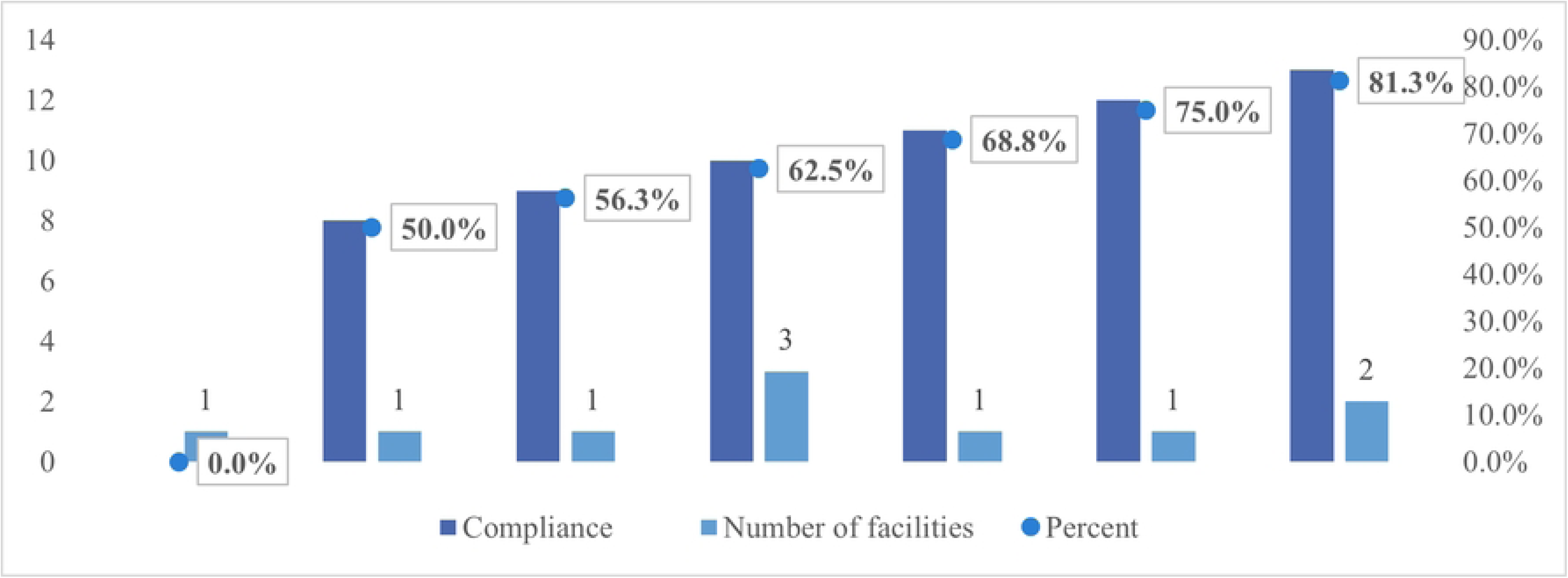
Compliance score.

### Service Integration, Documentation, Follow-Up, And Health Promotion Materials

The assessment revealed varying levels of service integration and support systems for cervical cancer prevention across the 10 facilities. Integration of cervical cancer screening with other services was generally high; 8 (80%) facilities had PITC/ CTC and STI services integration, 7 (70%) facilities had integrated family planning and maternal services. 9 (90%) facilities integrated cervical cancer screening with sexual and reproductive health (SRH) services. Inpatient service integration and the availability of integration guidelines were both reported in 6 (60%) facilities. Regarding documentation, 9 (90%) facilities-maintained registers/ logbooks, although data discrepancies were found in 2 (20%) facilities.

For follow-up mechanisms, 6 (60%) facilities had systems for cryotherapy follow-up, with only 3 (30%) facilities having systems for thermoablation follow-up. Referral follow-up mechanisms were present in 7 (70%) facilities. Client cards were used in 9 (90%) facilities, and IEC materials were available in 6 (60%) facilities. Community outreach for cervical cancer education was reported in 4 (40%), while 8 (80%) facilities conduct facility-based education (Figure 7). These findings reflect strong integration of cervical cancer services with other healthcare services and adequate documentation in most facilities, but notable gaps exist in patient’s follow-up, community education and thermoablation follow-up mechanisms.

**Fig 7:**
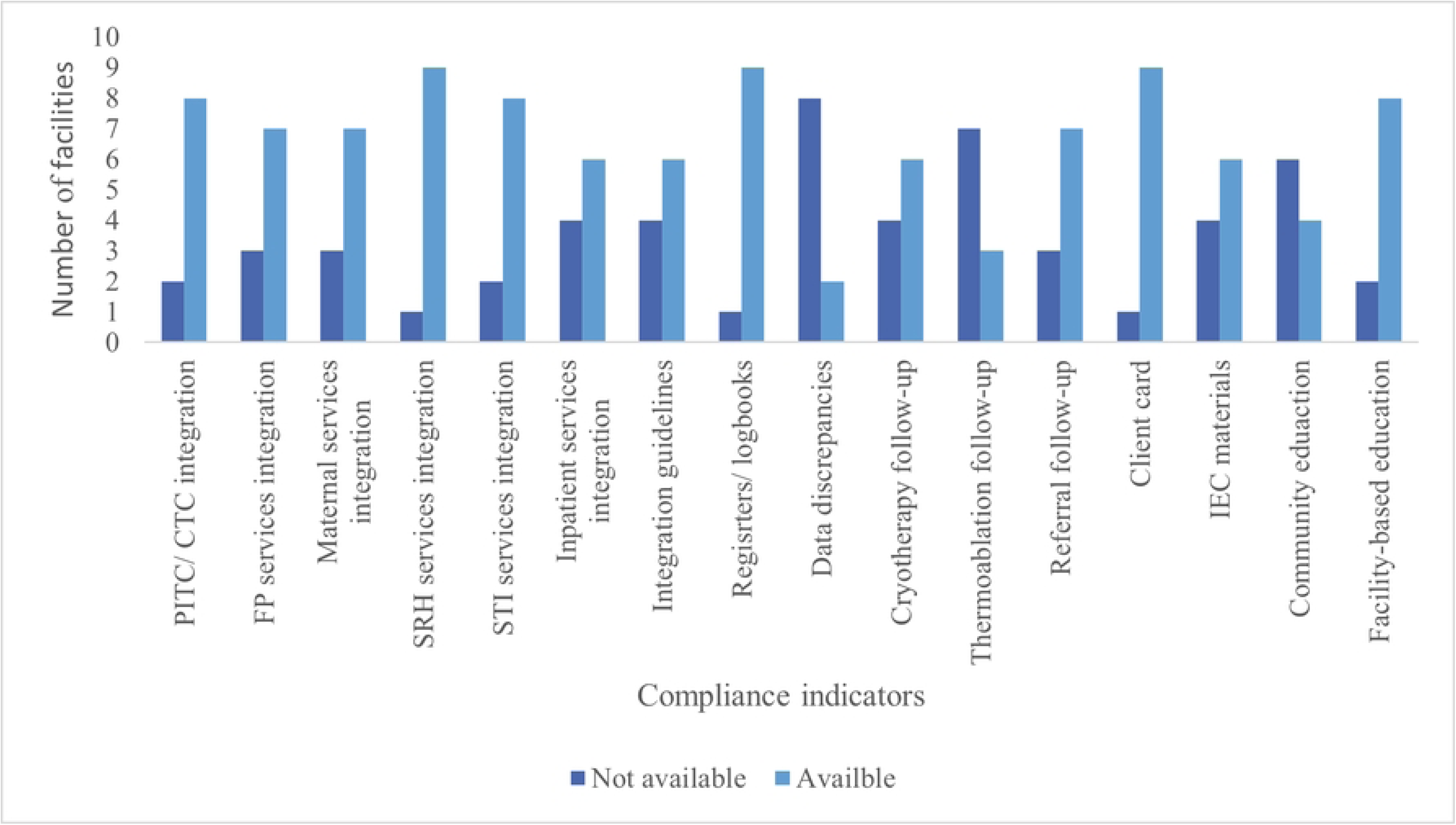
Compliance indicators.

### Compliance By Facility Levels

Compliance with the national guidelines for cervical cancer screening varied across facility levels. Referral hospitals recorded the highest compliance score of 81.3%, followed by district-level hospitals at 77.1% with both meeting the recommended compliance threshold of ≥70%. While health centers and dispensaries scored lower at 46.9% and 56.3% respectively (Fig 8).

**Fig 8:**
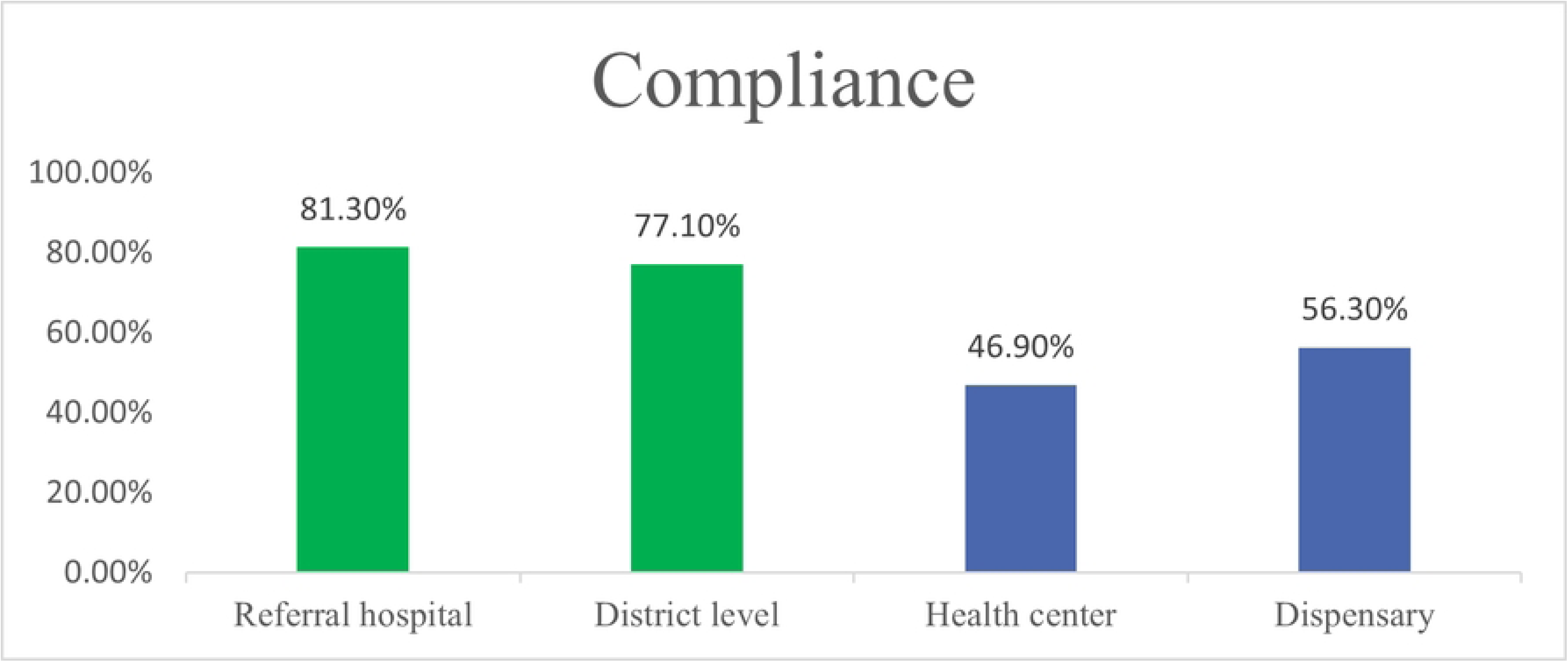
Compliance by facility level.

## Discussion

This study aimed to assess health facility readiness and guideline compliance for cervical cancer screening under the 2020–2024 national strategy in Kilimanjaro region. The key findings reveals that over half of the facilities were structurally ready, but the key gaps existed in the shortage of treatment equipment and qualified providers. Only one-third of Health Facility complied with national screening guidelines, with most facilities lacking proper follow-up systems and community outreach.

The assessment of structural readiness revealed that 60% of the assessed facilities met minimum readiness for cervical cancer secondary prevention services in the Kilimanjaro region, while 40% offered low-quality services based on CECAP scorecard. This highlight both limited availability and weak readiness, undermining the effectiveness of secondary prevention strategy. This evaluation also revealed that referral hospitals (tertiary level hospitals) are well structured and prepared for secondary prevention for cervical cancer compared to lower level hospital and this is similar to a study conducted in Bangladesh that found tertiary and specialized hospitals met 70% readiness index cutoff, while most other facilities lacked readiness, the results highlighted significant gap in CCS preparedness particularly at primary and secondary level (16). Key structural gaps included inadequate access to treatment equipment including 50% facilities had cryotherapy and 30% had thermoablation as well as limited private spaces, and shortage of trained personnel. Only 40% of facilities had staff trained in both VIA and cryotherapy, and 30% in VIA and thermoablation. These deficiencies directly affect the availability to delivery immediate treatment which is critical for cervical cancer progression (21). Compared to national and regional benchmarks, Kilimanjaro is lagging. Similar readiness challenges were reported by (14) in Tanzania and by (11) across SSA. In contrast, Rwanda and Burundi have shown that with strategic investment and centralized coordination, higher readiness and service quality are achievable (6).

The assessment of compliance to national guidelines for cervical cancer screening revealed that three quarter of facilities demonstrated low compliance with only a third quarter met the required 70% threshold on CECAP scorecard. Scores ranged from 0% to 81.3%, with one facility scoring zero indicating a complete lack of strategy implementation. These results highlight significant inconsistencies in applying national standards and raise concerns and raise concerns about service quality. These findings are consistence with a systematic review of cervical cancer screening in low-resource settings where screening guidelines exist in more than 80% of countries, adherence is variable, with many programs lacking provider compliance. The study found that supply gaps, and a lack of monitoring methods impede adherence, particularly in LMICs (15). According to the Tanzania Cervical Cancer Prevention and Control Plan 2020-2024, the target was to increase the proportion of health facilities offering high-quality CECAP services from 9% to 50% by 2024 (10). The findings suggest that Kilimanjaro is still behind in meeting this target, as even among the few facilities offering screening, most fall short in implementing standardized protocols. While services integration was strong, with 90% integrating services in SRH and 80% into PITC/ CTC services, gaps persist in follow-up systems, especially for thermoablation (30%) and community outreach education, conducted by only 40% of facilities. Although 90% had documentation systems, data discrepancies in 20% point to weak monitoring. These findings align with studies by Mugassa *et al.* (22) which show that poor compliance stems from weak supervision, lack of providers, training and inadequate tools. They also reinforce WHO’s concerns that availability alone is insufficient without adherence to clinical standards (21).

### Study strength and limitation

The study main strength is use of locally adapted, policy-aligned instruments which is modified CECAP scorecard tailored to Kilimanjaro’s healthcare environment. These instruments integrate regional referral pathways as well as national recommendations, ensuring that the findings are immediately relevant to local decision-makers.

This study faced various limitation: Poor record-keeping at some sites undermined the trustworthiness of service delivery data. Also, small sample size specifically for facility assessment limits generalizability to other regions or Tanzania’s national context.

## Conclusion

These findings suggest that the Kilimanjaro region is falling behind the strategic national goal of expanding the number of facilities providing high-quality CECAP services from 9% to 50% by 2024. The region had a very small proportion (8.9%) of all health institutions provided cervical cancer screening. Also, among this small proportion only 60% were structurally ready. The study also revealed that only 30% of the facilities that were providing cervical cancer screening complied fully with national guidelines for screening. While sexual and reproductive health and HIV services are well integrated in the region, there are still significant gaps of skilled providers, patient follow-up systems, and community education outreach.

The study emphasizes that simply having services available is insufficient; without structural readiness, compliance to clinical guidelines, and follow-up capacity, the impact of cervical cancer screening programs will be limited. These findings highlight the need for immediate action by health authorities and policy makers to address both systemic and operational hurdles that prevent Tanzania from fully achieving its cervical cancer prevention targets.

### Ethical Consideration

Ethical approval was obtained from the KCMC University Research and Ethical Review Committee (KURERC) with certificate number NO.PG11/25. Permission was obtained from the Regional Administrative Secretary (RAS) of the Kilimanjaro region and the respective hospitals. Participants were fully informed about the study’s purpose, and their participation was voluntary, with the option to withdraw at any time without explanation. All participants signed a written consent form prior to the study. A unique identification number was used instead of participants’ names to ensure confidentiality, and their identity was anonymous. The information gathered from participants was used only for research purposes and kept confidential.

## Data Availability

Data will be available at KCMC University library

## Acknowledgements

We are grateful to KCRI and NORA project for their generous support in this study. Also special thanks to the School of Public Health, department of community health at KCMC University for their support throughout this study.

## Author Contribution

**Conceptualization:** Paschal Yuda, Joackim Kessy, Gasper Mpehongwa and Declare L. Mushi

**Data curation:** Paschal Yuda, Anthony Kavindi

**Methodology:** Paschal Yuda, Anthony Kavindi, Gasper Mpehongwa, Joackim Kessy, Declare L. Mushi

**Formal analysis:** Paschal Yuda, Anthony Kavindi

**Writing original draft**: Paschal Yuda

**Writing- review and editing**: Paschal Yuda, Gasper Mpehongwa Joackim Kessy, Declare L. Mushi

